# Cohort study of Covid-19 vaccine effectiveness among healthcare workers in Finland, December 2020 - October 2021

**DOI:** 10.1101/2021.11.03.21265791

**Authors:** Eero Poukka, Ulrike Baum, Arto A. Palmu, Toni O. Lehtonen, Heini Salo, Hanna Nohynek, Tuija Leino

## Abstract

Recently, Covid-19 vaccine effectiveness has decreased especially against mild disease due to emergence of the Delta variant and waning protection. In this register-based study among healthcare workers in Finland, the vaccine effectiveness of two-dose mRNA vaccine series against SARS-CoV-2 infection decreased from 82% (95% CI 79-85%) 14-90 days after vaccination to 53% (43-62%) after 6 months. Similar trend was observed for other series. Waning was not observed against Covid-19 hospitalization. These results facilitate decision-making of booster doses for healthcare workers.

## Background

Covid-19 vaccines have been highly efficacious [1, 2]. However, their effectiveness has recently decreased, presumably for two reasons. First, the Delta variant, a strain capable of evading vaccine-induced immunity, emerged in spring 2021 [3-5]. Second, vaccine-induced immunity is waning [4-7]. Therefore, booster programs have been initiated in many countries [8, 9].

To facilitate decision-making concerning boosters for social and healthcare workers (HCW), we estimated vaccine effectiveness (VE) after the second dose. The objective was to evaluate whether protection declines after two doses of either mRNA or adenovirus vector (AdV) vaccines and after vaccination with first AdV and second mRNA vaccine (heterologous series).

### Vaccinations among healthcare workers in Finland

In Finland, the vaccination campaign started 27 December 2020 with vaccination of HCWs caring for Covid-19 patients and of personnel and residents in long-term care units [10]. They were vaccinated with mRNA vaccines (Comirnaty and Spikevax) using standard 3-4 weeks dosing interval through week 5, 2021. Then, simultaneously with introduction of AdV vaccine Vaxzevria, the dosing interval was extended to 12 weeks (Figure 1).

**Figure 1.**
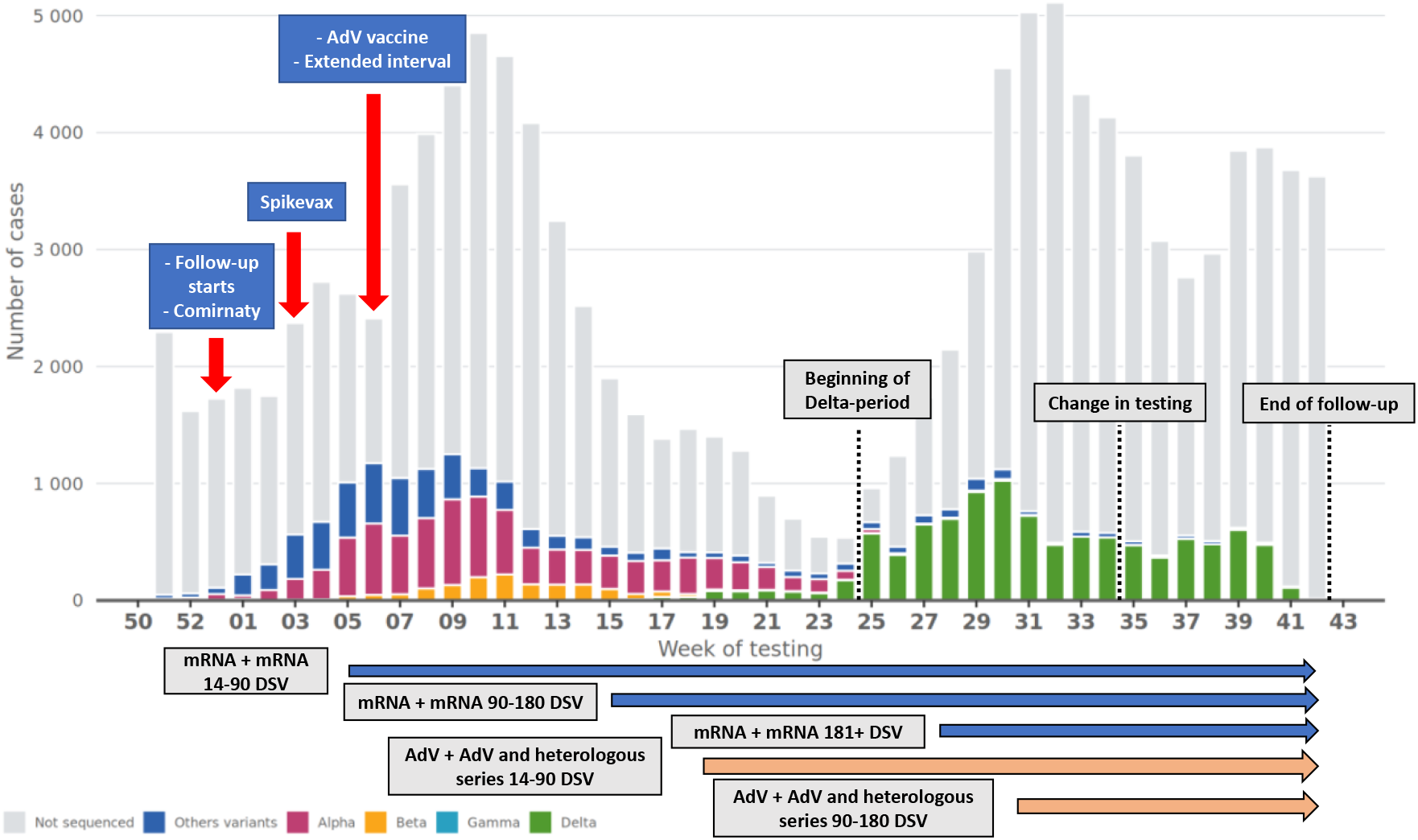
Number of laboratory-confirmed SARS-CoV-2 infections by variant if sequenced and calendar week. The horizontal arrows indicate the follow-up periods of two-dose series. Hospital districts ceased testing vaccinated persons with mild Covid-19-related symptoms 26 August 2021. DSV = Days since vaccination, AdV = Adenovirus vector.

### Register-based cohort study design

We conducted a nation-wide register-based cohort study to estimate the effectiveness of Covid-19 vaccines among 16–69-year-old HCWs in analogy to our previous study in elderly and chronically ill people [11]. Data of all people licensed to work as HCWs in Finland were extracted from the Social and Healthcare Professionals Register. HCWs included those in non-clinical positions because information on current position was unavailable. HCWs with laboratory-confirmed SARS-CoV-2 infection prior to 27 December 2020 were excluded.

The exposure was time since latest (first or second) Covid-19 vaccination recorded in the Finnish Vaccination Register. The study outcomes were laboratory-confirmed SARS-CoV-2 infection reported to the National Infectious Diseases Register and Covid-19-related hospitalization reported to the Care Register for Health Care in the week before or the two weeks after laboratory-confirmed SARS-CoV-2 infection [11].

### Statistical analysis

The follow-up started 27 December 2020. Each study subject was considered at risk of each outcome until first occurrence of any of the following events: outcome of interest, death, day 14 after confirmed infection, booster vaccination, or end of study (26 August if outcome was infection, 26 October 2021 otherwise). Using Cox regression, we compared the hazard of each outcome in vaccinated subjects with the corresponding hazard in the unvaccinated. The effect measure was VE, quantified as 1 minus the hazard ratio adjusted for age, sex, presence of medical conditions predisposing to severe Covid-19 (Supplementary Table 1), and residence in the most affected hospital district Helsinki-Uusimaa.

**Table 1.**
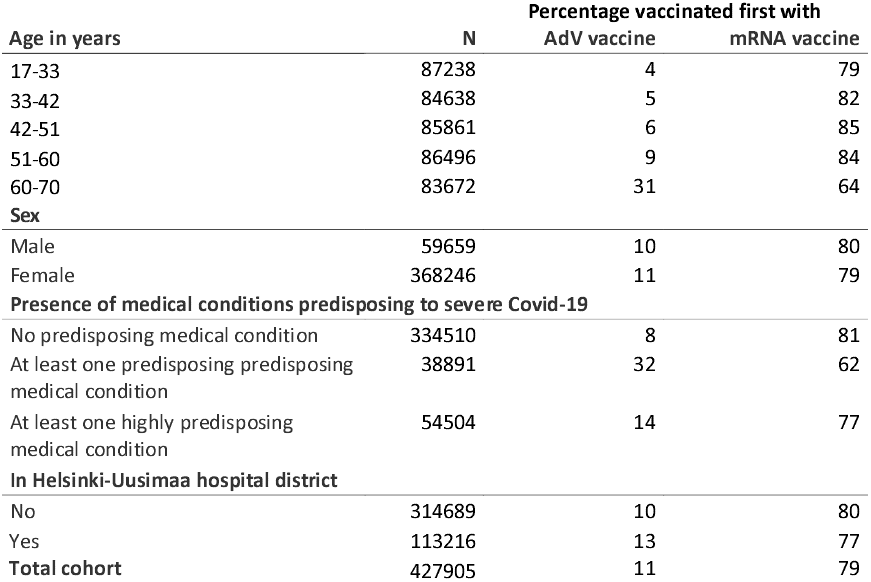
Distribution of baseline characteristics and percentage of study subjects presented by the first vaccine given. Healthcare workers (N = 427 905) in Finland, 27 Dec 2020 – 26 Oct 2021. AdV = Adenovirus vector.

| Age in years | N | Percentage vaccinated first with |  |
| --- | --- | --- | --- |
|  |  | AdV vaccine | mRNA vaccine |
| 17-33 | 87238 | 4 | 79 |
| 33-42 | 84638 | 5 | 82 |
| 42-51 | 85861 | 6 | 85 |
| 51-60 | 86496 | 9 | 84 |
| 60-70 | 83672 | 31 | 64 |
| **Sex** |  |  |  |
| Male | 59659 | 10 | 80 |
| Female | 368246 | 11 | 79 |
| **Presence of medical conditions predisposing to severe Covid-19** |  |  |  |
| No predisposing medical condition | 334510 | 8 | 81 |
| At least one predisposing medical condition | 38891 | 32 | 62 |
| At least one highly predisposing medical condition | 54504 | 14 | 77 |
| **In Helsinki-Uusimaa hospital district** |  |  |  |
| No | 314689 | 10 | 80 |
| Yes | 113216 | 13 | 77 |
| **Total cohort** | **427905** | **11** | **79** |

To estimate VE before and after emergence of the Delta variant, we split the follow-up on 21 June 2021 into pre-Delta and Delta period as Delta has accounted for the majority of sequenced cases since then (Figure 1). We also estimated brand-specific effectiveness of mRNA vaccines excluding those vaccinated first with AdV vaccine.

### Ethical concern

The study was conducted under the Finnish Communicable Disease Act and is part of the Finnish Institute for Health and Welfare surveillance duty to monitor the effectiveness of vaccines [12]. Therefore, the study did not require further ethical review.

### Description of the study cohort

The cohort included 427905 HCWs: 291758 (68%) nurses, 23886 (6%) physicians, 13074 (3%) dentists and oral hygienists and 99187 (23%) other professionals. By the end of follow-up 90% of the HCWs had received at least one vaccination. Two-dose series of mRNA vaccines were received by 315413 (74%) HCWs. Two-dose AdV vaccine and heterologous series were administered to 14760 (3%) and 30548 (7%) HCWs.

### Estimates of vaccine effectiveness

There were 3874 and 1757 laboratory-confirmed SARS-CoV-2 infections in the unvaccinated and vaccinated. VE against infection was 82% (95% confidence interval 79-85%) for mRNA, 89% (73-95%) for AdV and 80% (72-86%) for heterologous vaccine series 14-90 days after the second dose. However, VE appeared to wane over time (Figure 2, Supplementary Table 2). VE was 62% (55-68%) for mRNA, 63% (−166-95%) for AdV and 62% (30-79%) for heterologous vaccine series 91-180 days after the second dose.

**Figure 2.**
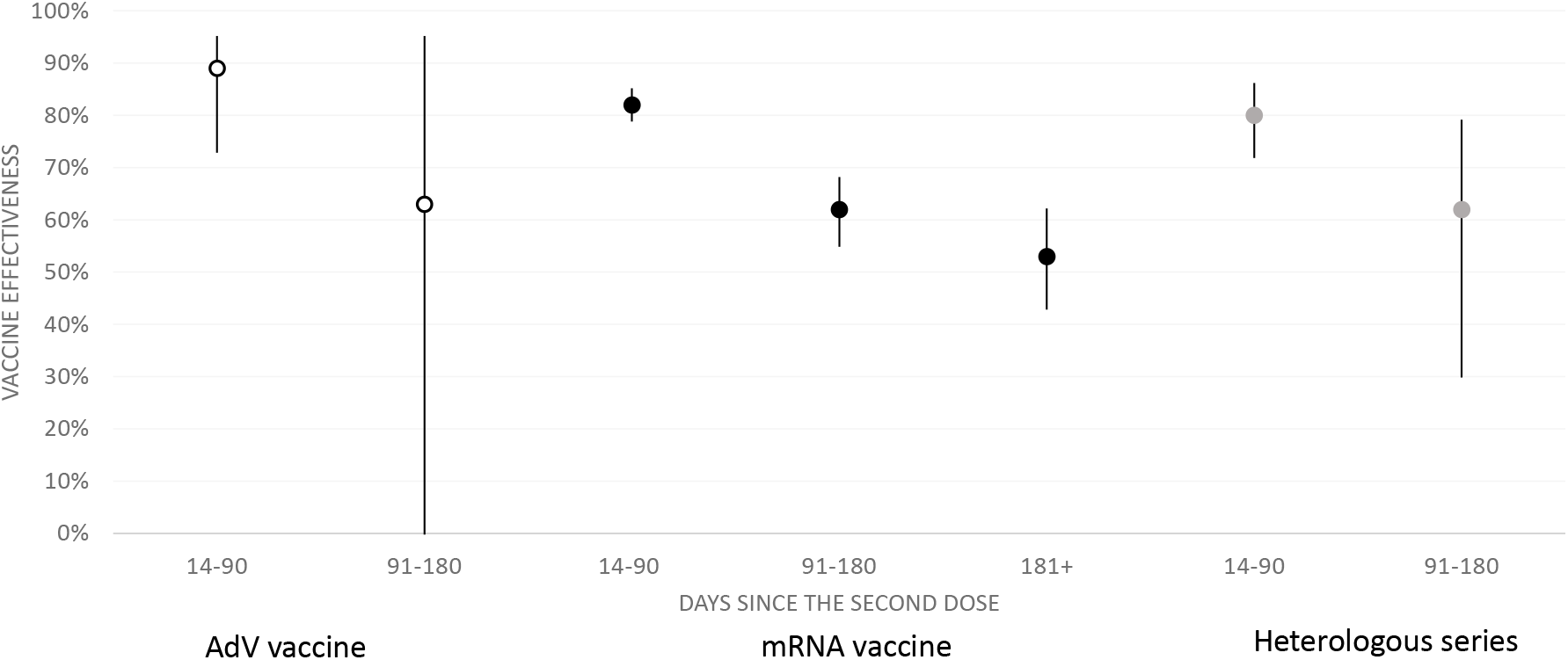
Effectiveness of AdV (white), mRNA (black) and heterologous (grey) vaccine series against laboratory-confirmed SARS-CoV-2 infection among healthcare workers (N = 427 905) in Finland, 27 Dec 2020 – 26 Aug 2021. AdV = Adenovirus vector.

There were 220 and 35 Covid-19-related hospitalizations in the unvaccinated and vaccinated. Effectiveness against hospitalization was 88% or above for all series during the first ten months of the campaign (Figure 3, Supplementary Table 3).

**Figure 3.**
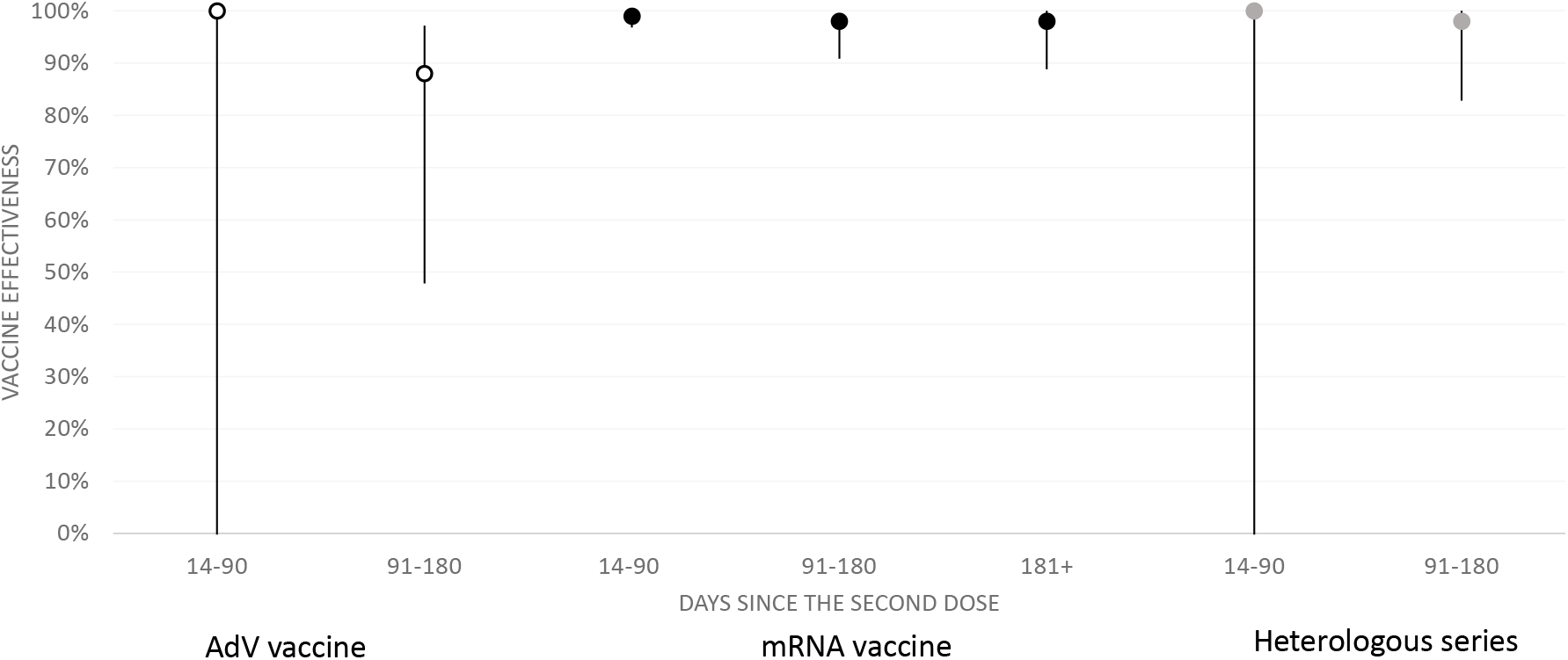
Effectiveness of AdV (white), mRNA (black) and heterologous (grey) vaccine series against Covid-19-related hospitalization among healthcare workers (N = 427 905) in Finland, 27 Dec 2020 – 26 Oct 2021.

During the Delta period VE was similar to that in the Pre-Delta period (Supplementary Tables 2-3). Brand-specific mRNA VE estimates were also comparable (Supplementary Table 4).

## Discussion

This study’s aim was to analyze the effectiveness of Covid-19 vaccines in HCWs during the first 10 months of the vaccination campaign in Finland. We observed high early VE against SARS-CoV-2 infection after the second dose with significant waning after 3 months. However, Covid-19 vaccines maintained excellent effectiveness against Covid-19 hospitalization. We found no meaningful differences between mRNA and AdV vaccine series, although current knowledge is that mRNA vaccine provides better protection [4, 13]. As in Denmark and Sweden, we observed similar protection levels after mRNA and heterologous vaccine series [14, 15].

We did not detect changes in VE following the emergence of Delta, indicating that the decrease in VE against infection is due to waning of vaccine-induced immunity, which has also been seen elsewhere [4, 6, 7, 13, 16]. The waning against infection was slightly faster than observed in a clinical trial of Comirnaty [7] and among HCWs in USA [16], but comparable to the waning detected among 18–64 year-old people in UK [4]. Because booster doses enhance protection against SARS-CoV-2 infections among 16 years and older [8], they could be beneficial to HCWs. Covid-19 vaccines might also reduce transmission of SARS-CoV-2 [17] and, therefore, boosters among HCWs could indirectly protect patients.

Although in agreement with the literature, our findings may be prone to bias. The HCWs with the longest post-vaccination follow-up are those at high risk of work-related infection, such as intensive care unit nurses. Therefore, our study may underestimate the average VE in HCWs 6 months after the second dose. Furthermore, the HCWs who were vaccinated first received their second dose 3-4 weeks after the first one, while those vaccinated later received their second dose 12 weeks after the first one. Thus, the length of follow-up after receipt of the second dose differs between these groups. As it has been shown that, compared to the standard interval, an extended dosing interval induces greater antibody response and possibly greater VE [18-20], this could have introduced further underestimation of the average VE 6 months after the second dose.

Strengths of this analysis are the size and representativeness of the cohort and the register-based nature of the study. Unfortunately, the study data did not include information on the HCWs’ current status and field of employment so it was impossible to assess their risk of work-related infection.

## Conclusions

We observed waning of Covid-19 VE against SARS-CoV-2 infection three and six months after the second dose, while high VE against hospitalization sustained beyond six months. Boosters may be beneficial for HCWs to enhance protection against infection and to decrease transmission of SARS-CoV-2 to patients. However, level and duration of protection after a booster dose are presently unknown.

## Supporting information

Ethical concern

## Data Availability

By Finnish law, the authors are not permitted to share individual-level register data. The computing code is available upon request.

## Authors’ contributions

EP, UB, TL, HN and AAP conceptualized the study. HS and TOL identified the medical conditions predisposing to severe Covid-19. UB conducted the statistical analysis. EP reviewed the literature and drafted the manuscript. All authors gave comments and revised the manuscript.

## Acknowledgements

The authors thank Esa Ruokokoski, Jonas Sundman, Oskari Luomala, Teemu Möttönen and Tuomo Nieminen from the Finnish Institute for Health and Welfare for their assistance in managing the study data.

