## Supplement material for "Cohort study of Covid-19 vaccine effectiveness among healthcare workers in Finland, December 2020 - October 2021"

**Supplementary Table 1:** Definition of medical conditions (highly) predisposing to severe Covid-19.

| Medical condition | Classification | Codes | Register |
|---|---|---|---|
| **Highly predisposing to severe Covid-19** | | | |
| Organ or stem cell transplant | ICD-10 | T86, Z94 | 1,2 |
| Active cancer treatment | ICD-10 | C00–C97 (except C44), D05.1, D39 | 1,2 |
| Severe disorders of the immune system | ICD-10 | D70.8, D80–D84, E31.00 | 2 |
| Severe chronic renal disease | ICD-10 | I12, I13, N00–N05, N07, N08, N11, N14, N18, N19, E10.2, E11.2, E14.2 | 1,2 |
| Asthma requiring continuous medication | ICD-10 | J45, J46 | 2, 3 |
| | ICPC-2 | R96 | 3 |
| Severe chronic pulmonary disease | ICD-10 | J41–J44, J47, Z90.2 | 2 |
| Type 2 diabetes mellitus requiring medication | ICD-10 | E11, E13, E14 | 2,3 |
| | ICPC-2 | T90 | 3 |
| Blood glucose lowering drugs, excluding insulin | ATC | A10B | 4 |
| Down syndrome | ICD-10 | Q90 | 2, 3 |
| **Predisposing to severe Covid-19** | | | |
| Severe heart disease | ICD-10 | I11–I13, I15, I20–I25, I50 | 1, 2 |
| Neurological illness or condition that affects breathing | ICD-10 | G70-G73, G80–G83, I60–I69 | 2 |
| Immunosuppressive drug therapy for autoimmune disease | | | |
| Autoimmune disease | ICD-10 | D86, K50, K51, L40, M02, M05–M07, M13.9, M45, M46.0, M46.1, M46.9, M94.1 | 1, 2 |
| Immunosuppressive drug therapy | ATC | H02AB02, H02AB04, H02AB06, H02AB07, L01BA01, L01XC02, L04AA06, L04AA10, L04AA13, L04AA18, L04AA24, L04AA26, L04AA29, L04AA33, L04AA37, L04AB, L04AC, L04AD01, L04AD02, L04AX01, L04AX03 | 4 |
| Severe chronic liver disease | ICD-10 | K70.2, K70.3, K70.4, K71–K74 | 2 |
| Type 1 diabetes | ICD-10 | E10 | 2, 3 |
| | ICPC-2 | T89 | 3 |
| Insulin and analogues | ATC | A10A | 4 |
| Adrenal insufficiency | ICD-10 | E25.0, E27.1, E27.2, E27.4, E31.00, E31.01, E31.08, E89.6 | 1, 2 |
| Sleep apnea | ICD-10 | G47.3 | 2, 3 |
| Continuous positive airway pressure therapy | NCSP | WX723, WX780 | 2 |
| Psychotic disorders | ICD-10 | F20–F29 | 2, 3 |
| | ICPC-2 | P72 | 3 |
| Clozapine | ATC | N05AH02 | 4 |

ATC, Anatomical Therapeutic Chemical Classification System; ICD-10, International Statistical Classification of Diseases and Related Health Problems, tenth revision; ICPC-2, International Classification of Primary Care, second edition; NCSP, Nordic Nomesco Classification of Surgical Procedures.

Registers: 1, Special Reimbursement Register for Medicine Expenses; 2, Care Register for Health Care; 3, Register of Primary Health Care Visits; 4, Prescription Centre database.

**Supplementary Table 2:** Vaccine effectiveness against laboratory-confirmed SARS-CoV-2 infection in healthcare workers.

| | Main analysis | | | | | Sensitivity analysis | | | | | | | | | |
|---|---|---|---|---|---|---|---|---|---|---|---|---|---|---|---|
| | 27 Dec 2020 – 26 Aug 2021 | | | | | 27 Dec 2020 – 20 Jun 2021 (Pre-Delta) | | | | | 21 Jun 2021 – 26 Aug 2021 (Delta) | | | | |
| | Cases | AR | VE | CI95% | | Cases | AR | VE | CI95% | | Cases | AR | VE | CI95% | |
| **Not vaccinated** | 3874 | 2066 | Reference | | | 3287 | 1086 | Reference | | | 587 | 991 | Reference | | |
| **AdV vaccine (0-20 DSV)** | 43 | 310 | 36 | 14 | 53 | 43 | 310 | 36 | 13 | 53 | 0 | 0 | 100 | -Inf | 100 |
| **AdV vaccine (21-41 DSV)** | 48 | 402 | 7 | -24 | 30 | 48 | 402 | 6 | -26 | 29 | 0 | 0 | 100 | -Inf | 100 |
| **AdV vaccine (42+ DSV)** | 50 | 7144 | 22 | -3 | 42 | 44 | 6877 | 15 | -15 | 37 | 6 | 287 | 49 | -16 | 77 |
| **AdV vaccine + AdV vaccine (0-13 DSV)** | < 5 | 112 | 65 | -40 | 91 | < 5 | 88 | -16 | -727 | 84 | < 5 | 25 | 79 | -51 | 97 |
| **AdV vaccine + AdV vaccine (14-90 DSV)** | 5 | 37 | 89 | 73 | 95 | 0 | 0 | 100 | -Inf | 100 | 5 | 37 | 88 | 71 | 95 |
| **AdV vaccine + AdV vaccine (91-180 DSV)** | < 5 | 38 | 63 | -166 | 95 | 0 | 0 | 100 | -Inf | 100 | < 5 | 38 | 62 | -177 | 95 |
| **AdV vaccine + AdV vaccine (181+ DSV)** | - | - | - | - | - | - | - | - | - | - | - | - | - | - | - |
| **AdV vaccine + mRNA vaccine (0-13 DSV)** | 7 | 100 | 43 | -20 | 73 | 6 | 64 | 27 | -65 | 67 | < 5 | 36 | 73 | -95 | 96 |
| **AdV vaccine + mRNA vaccine (14-90 DSV)** | 38 | 144 | 80 | 72 | 86 | 0 | 0 | 100 | -Inf | 100 | 38 | 144 | 80 | 72 | 86 |
| **AdV vaccine + mRNA vaccine (91-180 DSV)** | 11 | 180 | 62 | 30 | 79 | 0 | 0 | 100 | 100 | 100 | 11 | 180 | 63 | 33 | 80 |
| **AdV vaccine + mRNA vaccine (181+ DSV)** | - | - | - | - | - | - | - | - | - | - | - | - | - | - | - |
| **mRNA vaccine (0-20 DSV)** | 283 | 1779 | 22 | 11 | 31 | 208 | 1187 | 10 | -4 | 22 | 75 | 599 | 46 | 31 | 57 |
| **mRNA vaccine (21-41 DSV)** | 184 | 1015 | 48 | 39 | 55 | 78 | 572 | 40 | 25 | 52 | 106 | 446 | 56 | 46 | 64 |
| **mRNA vaccine (42+ DSV)** | 542 | 956 | 40 | 34 | 46 | 87 | 458 | 38 | 23 | 50 | 455 | 501 | 45 | 37 | 51 |
| **mRNA vaccine + mRNA vaccine (0-13 DSV)** | 110 | 422 | 70 | 64 | 76 | 21 | 177 | 70 | 54 | 81 | 89 | 245 | 71 | 64 | 77 |
| **mRNA vaccine + mRNA vaccine (14-90 DSV)** | 162 | 326 | 82 | 79 | 85 | 80 | 209 | 77 | 71 | 82 | 82 | 118 | 85 | 81 | 88 |
| **mRNA vaccine + mRNA vaccine (91-180 DSV)** | 168 | 377 | 62 | 55 | 68 | 28 | 100 | 55 | 34 | 69 | 140 | 277 | 65 | 58 | 71 |
| **mRNA vaccine + mRNA vaccine (181+ DSV)** | 103 | 320 | 53 | 43 | 62 | - | - | - | - | - | 103 | 320 | 56 | 46 | 65 |

AR, Attack rate (cumulative risk multiplied by $10^5$); CI, Confidence interval; DSV, Days since vaccination; VE, Vaccine effectiveness.

**Supplementary Table 3:** Vaccine effectiveness against Covid-19 related hospitalization in healthcare workers.

| | Main analysis | | | | | Sensitivity analysis | | | | | | | | | |
|---|---|---|---|---|---|---|---|---|---|---|---|---|---|---|---|
| | 27 Dec 2020 – 26 Oct 2021 | | | | | 27 Dec 2020 – 20 Jun 2021 (Pre-Delta) | | | | | 21 Jun 2021 – 26 Oct 2021 (Delta) | | | | |
| | Cases | AR | VE | CI95% | | Cases | AR | VE | CI95% | | Cases | AR | VE | CI95% | |
| **Not vaccinated** | 220 | 198 | Reference | | | 149 | 51 | Reference | | | 71 | 147 | Reference | | |
| **AdV vaccine (0-20 DSV)** | < 5 | 11 | 69 | -26 | 92 | < 5 | 11 | 70 | -22 | 93 | 0 | 0 | 100 | -Inf | 100 |
| **AdV vaccine (21-41 DSV)** | 7 | 45 | -17 | -156 | 47 | 7 | 45 | -13 | -149 | 49 | 0 | 0 | 100 | -Inf | 100 |
| **AdV vaccine (42+ DSV)** | < 5 | 52 | 88 | 10 | 98 | 0 | 0 | 100 | -Inf | 100 | < 5 | 52 | 42 | -330 | 92 |
| **AdV vaccine + AdV vaccine (0-13 DSV)** | 0 | 0 | 100 | -Inf | 100 | 0 | 0 | 100 | -Inf | 100 | 0 | 0 | 100 | -Inf | 100 |
| **AdV vaccine + AdV vaccine (14-90 DSV)** | 0 | 0 | 100 | -Inf | 100 | 0 | 0 | 100 | -Inf | 100 | 0 | 0 | 100 | -Inf | 100 |
| **AdV vaccine + AdV vaccine (91-180 DSV)** | < 5 | 28 | 88 | 48 | 97 | - | - | - | - | - | < 5 | 28 | 81 | 9 | 96 |
| **AdV vaccine + AdV vaccine (181+ DSV)** | 0 | 0 | 100 | -Inf | 100 | - | - | - | - | - | 0 | 0 | 100 | -Inf | 100 |
| **AdV vaccine + mRNA vaccine (0-13 DSV)** | 0 | 0 | 100 | -Inf | 100 | 0 | 0 | 100 | -Inf | 100 | 0 | 0 | 100 | -Inf | 100 |
| **AdV vaccine + mRNA vaccine (14-90 DSV)** | 0 | 0 | 100 | -Inf | 100 | 0 | 0 | 100 | -Inf | 100 | 0 | 0 | 100 | -Inf | 100 |
| **AdV vaccine + mRNA vaccine (91-180 DSV)** | < 5 | 4 | 98 | 83 | 100 | - | - | - | - | - | < 5 | 4 | 97 | 81 | 100 |
| **AdV vaccine + mRNA vaccine (181+ DSV)** | 0 | 0 | 100 | -Inf | 100 | - | - | - | - | - | 0 | 0 | 100 | 100 | 100 |
| **mRNA vaccine (0-20 DSV)** | < 5 | 95 | 87 | 58 | 96 | < 5 | 95 | 79 | 31 | 93 | 0 | 0 | 100 | -Inf | 100 |
| **mRNA vaccine (21-41 DSV)** | < 5 | 2 | 95 | 67 | 99 | < 5 | 2 | 89 | 15 | 98 | 0 | 0 | 100 | -Inf | 100 |
| **mRNA vaccine (42+ DSV)** | 13 | 30 | 83 | 70 | 91 | < 5 | 5 | 90 | 27 | 99 | 12 | 25 | 83 | 68 | 91 |
| **mRNA vaccine + mRNA vaccine (0-13 DSV)** | 0 | 0 | 100 | -Inf | 100 | 0 | 0 | 100 | -Inf | 100 | 0 | 0 | 100 | -Inf | 100 |
| **mRNA vaccine + mRNA vaccine (14-90 DSV)** | < 5 | 2 | 99 | 97 | 100 | < 5 | 2 | 95 | 64 | 99 | < 5 | 1 | 100 | 97 | 100 |
| **mRNA vaccine + mRNA vaccine (91-180 DSV)** | < 5 | 3 | 98 | 91 | 99 | 0 | 0 | 100 | -Inf | 100 | < 5 | 3 | 98 | 90 | 99 |
| **mRNA vaccine + mRNA vaccine (181+ DSV)** | < 5 | 2 | 98 | 89 | 100 | - | - | - | - | - | < 5 | 2 | 98 | 88 | 100 |

AR, Attack rate (cumulative risk multiplied by $10^5$); CI, Confidence interval; DSV, Days since vaccination; VE, Vaccine effectiveness.

**Supplementary Table 4:** Brand-specific mRNA vaccine effectiveness against laboratory-confirmed SARS-COV-2 infection and Covid-19 related hospitalization in healthcare workers excluding those vaccinated first with AdV vaccine.

| | **SARS-CoV-2 infection** | | | | | **Covid-19 related hospitalization** | | | | |
|---|---|---|---|---|---|---|---|---|---|---|
| | 27 Dec 2020 – 26 Aug 2021 | | | | | 27 Dec 2020 – 26 Oct 2021 | | | | |
| | Cases | AR | VE | CI95% | | Cases | AR | VE | CI95% | |
| **Not vaccinated** | 3838 | 2141 | Reference | | | 218 | 201 | Reference | | |
| **Comirnaty (0-20 DSV)** | 265 | 1905 | 21 | 11 | 31 | < 5 | 111 | 85 | 54 | 95 |
| **Comirnaty (21-41 DSV)** | 174 | 1062 | 47 | 38 | 55 | < 5 | 3 | 95 | 63 | 99 |
| **Comirnaty (42+ DSV)** | 507 | 985 | 40 | 33 | 46 | 12 | 34 | 82 | 68 | 90 |
| **Comirnaty + Comirnaty (0-13 DSV)** | 104 | 441 | 71 | 64 | 76 | 0 | 0 | 100 | -Inf | 100 |
| **Comirnaty + Comirnaty (14-90 DSV)** | 154 | 323 | 83 | 80 | 85 | < 5 | 2 | 99 | 97 | 100 |
| **Comirnaty + Comirnaty (91-180 DSV)** | 167 | 377 | 63 | 56 | 69 | < 5 | 3 | 98 | 91 | 99 |
| **Comirnaty + Comirnaty (181+ DSV)** | 103 | 320 | 55 | 45 | 64 | < 5 | 2 | 98 | 89 | 100 |
| **Comirnaty + Spikevax (0-13 DSV)** | < 5 | 173 | 65 | -146 | 95 | 0 | 0 | 100 | -Inf | 100 |
| **Comirnaty + Spikevax (14-90 DSV)** | 0 | 0 | 100 | -Inf | 100 | 0 | 0 | 100 | -Inf | 100 |
| **Comirnaty + Spikevax (91-180 DSV)** | 0 | 0 | 100 | -Inf | 100 | 0 | 0 | 100 | -Inf | 100 |
| **Comirnaty + Spikevax (181+ DSV)** | 0 | 0 | 100 | -Inf | 100 | 0 | 0 | 100 | -Inf | 100 |
| **Spikevax (0-20 DSV)** | 18 | 551 | 55 | 29 | 72 | 0 | 0 | 100 | -Inf | 100 |
| **Spikevax (21-41 DSV)** | 10 | 354 | 72 | 47 | 85 | 0 | 0 | 100 | -Inf | 100 |
| **Spikevax (42+ DSV)** | 35 | 393 | 61 | 45 | 72 | < 5 | 7 | 89 | 22 | 98 |
| **Spikevax + Comirnaty (0-13 DSV)** | 0 | 0 | 100 | -Inf | 100 | 0 | 0 | 100 | -Inf | 100 |
| **Spikevax + Comirnaty (14-90 DSV)** | 0 | 0 | 100 | -Inf | 100 | 0 | 0 | 100 | -Inf | 100 |
| **Spikevax + Comirnaty (91-180 DSV)** | 0 | 0 | 100 | -Inf | 100 | 0 | 0 | 100 | -Inf | 100 |
| **Spikevax + Comirnaty (181+ DSV)** | 0 | 0 | 100 | -Inf | 100 | 0 | 0 | 100 | 100 | 100 |
| **Spikevax + Spikevax (0-13 DSV)** | 5 | 113 | 83 | 60 | 93 | 0 | 0 | 100 | -Inf | 100 |
| **Spikevax + Spikevax (14-90 DSV)** | 8 | 322 | 84 | 68 | 92 | 0 | 0 | 100 | -Inf | 100 |
| **Spikevax + Spikevax (91-180 DSV)** | < 5 | 179 | 69 | -124 | 96 | 0 | 0 | 100 | -Inf | 100 |
| **Spikevax + Spikevax (181+ DSV)** | - | - | - | - | - | 0 | 0 | 100 | -Inf | 100 |

AR, Attack rate (cumulative risk multiplied by $10^5$); CI, Confidence interval; DSV, Days since vaccination; VE, Vaccine effectiveness.

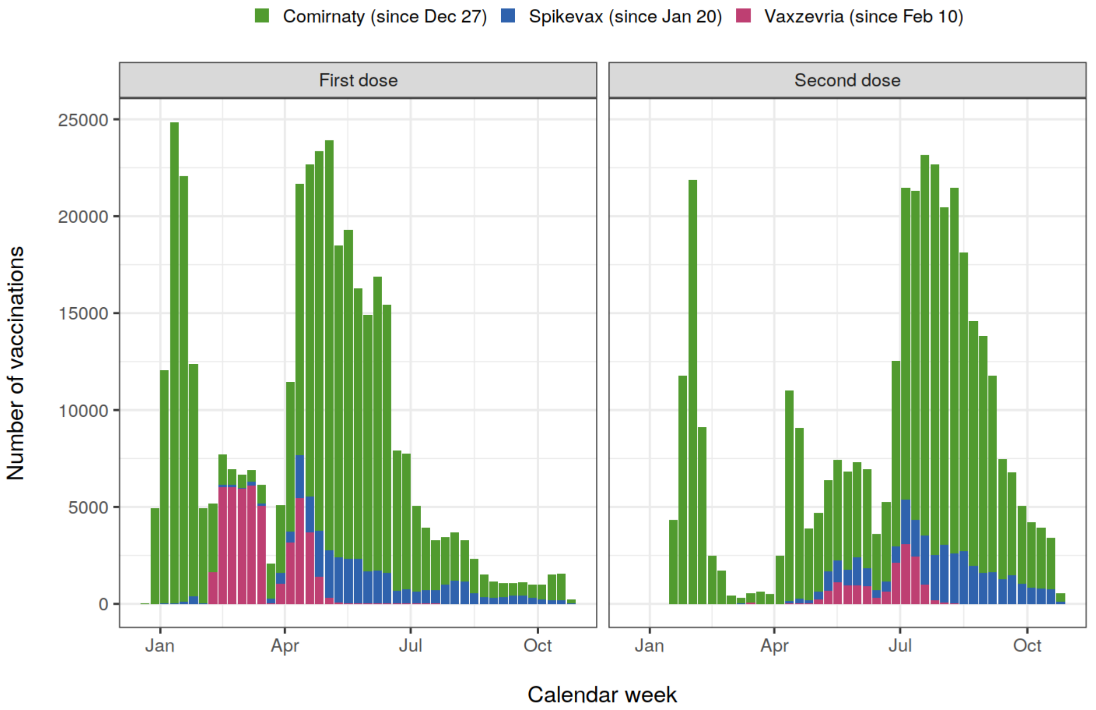


**Supplementary Figure 1.** Number of vaccinations administered in healthcare workers by calendar week.

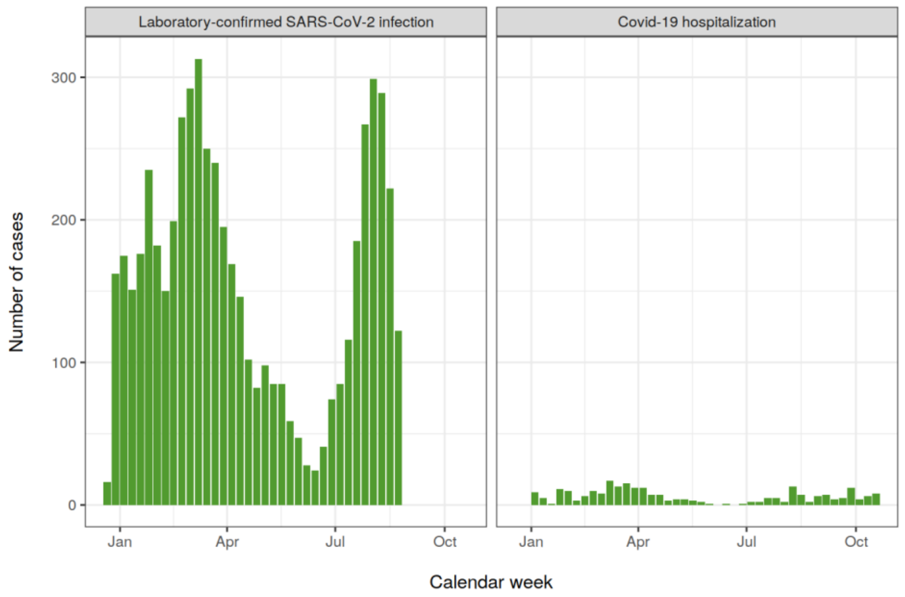


**Supplementary Figure 2:** Number of laboratory-confirmed SARS-CoV-2 infections and Covid-19 related hospitalizations in healthcare workers by calendar week.
