## Supplementary material for "Cohort study of Covid-19 vaccine effectiveness among healthcare workers in Finland, December 2020 - October 2021": Ethical concern

To whom it may concern:

As director of the department for Health security of the Finnish Institute for Health and Welfare, I certify that:

- I am the competent auhority for assessing whether research requires institutionnal ethical review or if the Finnish communicable diseases law (Tartuntatautilaki 1227/2016)and the law on the duties of the Finnish institute for Health (Laki Terveyden ja hyvinvoinnin laitoksesta 668/2008) and Welfare allows the implementation of the research without seeking further ethical review.
- The research presented by Poukka et al in "**Cohort study of Covid-19 vaccine effectiveness among healthcare workers in Finland, December 2020 - October 2021**" did not require further ethical review before implementation as its aim was to monitor vaccine effectiveness of infectious disease (Tartuntatautilaki 1227/2016).

Helsinki, October 29th 2021

Prof Mika Salminen
